# Development and Validation of Collaborative Robot-assisted Cutting Method for Iliac Crest Flap Raising: Randomized Crossover Trial

**DOI:** 10.1101/2024.09.04.24312594

**Authors:** Paulina Becker, Yao Li, Sergey Drobinsky, Jan Egger, Kunpeng Xie, Ashkan Rashad, Klaus Radermacher, Rainer Röhrig, Matías de la Fuente, Frank Hölzle, Behrus Puladi

**Affiliations:** Department of Oral and Maxillofacial Surgery, University Hospital RWTH Aachen, Aachen, Germany; Institute of Medical Informatics, University Hospital RWTH Aachen, Aachen, Germany; Chair of Medical Engineering, RWTH Aachen University, 52074 Aachen, Germany; Institute for Artificial Intelligence in Medicine, University of Duisburg-Essen, 45131 Essen, Germany

**Keywords:** Surgical robotics, Computer-assisted surgery, Iliac crest flap, DCIA flap, Cutting guide, Augmented Reality

## Abstract

The current gold standard of computer-assisted jaw reconstruction includes raising microvascular bone flaps with patient-specific 3D-printed cutting guides. The downsides of cutting guides are invasive fixation, periosteal denudation, preoperative lead time and missing intraoperative flexibility. This study aimed to investigate the feasibility and accuracy of a robot-assisted cutting method for raising iliac crest flaps compared to a conventional 3D-printed cutting guide.

In a randomized crossover design, 40 participants raised flaps on pelvic models using conventional cutting guides and a robot-assisted cutting method. The accuracy was measured and compared regarding osteotomy angle deviation, Hausdorff Distance (HD) and Average Hausdorff Distance (AVD). Duration, workload and usability were further evaluated.

The mean angular deviation for the robot-assisted cutting method was 1.9±1.1° (mean±sd) and for the 3D-printed cutting guide it was 4.7±2.9° (p<0.001). The HD resulted in a mean value of 1.5±0.6mm (robot) and 2.0±0.9mm (conventional) (p<0.001). For the AVD, this was 0.8±0.5mm (robot) and 0.8±0.4mm (conventional) (p=0.320). Collaborative robot-assisted cutting is an alternative to 3D-printed cutting guides in experimental static settings, achieving slot design benefits with less invasiveness and higher intraoperative flexibility. In the next step, the results should be tested in a dynamic environment with a moving phantom and on the cadaver.

## Introduction

Surgical reconstruction of the lower jaw (mandible) and upper jaw (maxilla) is a complex procedure in oral and maxillofacial surgery (OMFS), that is performed to restore bone continuity and its physiological functions, such as mastication, swallowing or speech [1]. In addition to functionality, the surgery must meet aesthetic requirements as well [2]. Common reasons for jaw discontinuity are tumors, congenital malformations, severe osteomyelitis/osteonecrosis or severe trauma [3]. The reconstruction of the jaw can be performed using different donor sites. Typically, vascularized flaps are raised from the fibula, iliac crest, or scapula, each having its strengths and limitations [4].

In recent years, computer-assisted surgery (CAS) has become the gold standard for maxillofacial reconstruction compared to freehand reconstruction. Based on surface models from preoperative computed tomography (CT) or cone beam computed tomography (CBCT) scans, CAS involves virtual surgical planning (VSP) of the lower or upper jaw resection and corresponding bone reconstruction with an osseous flap. The preoperative plan is then translated to the operating room using 3D-printed cutting guides for both jaw resection and flap raising [5]. This increases the accuracy and safety of bone resection, including flap raising, while decreasing surgical time and duration of ischemia [6].

However, 3D-printed guides have several downsides: A lack of intraoperative flexibility due to the need of preoperative design, fabrication, and sterilization [7]; the manufacturing process itself is time-consuming and costly [6]; the need of operative invasive fixation of the guide, including some periosteal denudation to ensure proper placement of the guide, which can potentially compromise bone perfusion and could cause osteonecrosis [8–10];

For this reason, an attempt was already made in 2011 to use classic navigation instead of 3D-printed cutting guides [11,12]. However, surgical navigation has the disadvantage that the spatial separation between the surgical field and the surgical navigation has a negative impact on hand-eye coordination and depth perception [12,13], which worsens with increasing complexity of the surgical task. Therefore, several studies have attempted to develop alternative flap raising systems with robotic approaches [7,14–18] as well as augmented reality (AR) [19–23] (Table 1). Both methods have different advantages and disadvantages. So far, robotic approaches have only been investigated for free fibula flaps (FFF) and on the mandibula [24], but no studies investigated haptic robot-assisted methods for deep circumflex iliac artery (DCIA) flap harvesting. However, the results about FFF harvesting are not directly transferable to DCIA raising because unlike FFF, where only isolated vertical osteotomies are required due to the anatomy of the fibula, the anatomy of the iliac crest requires at least one horizontal osteotomy to connect the osteotomy planes.

**Table 1.**
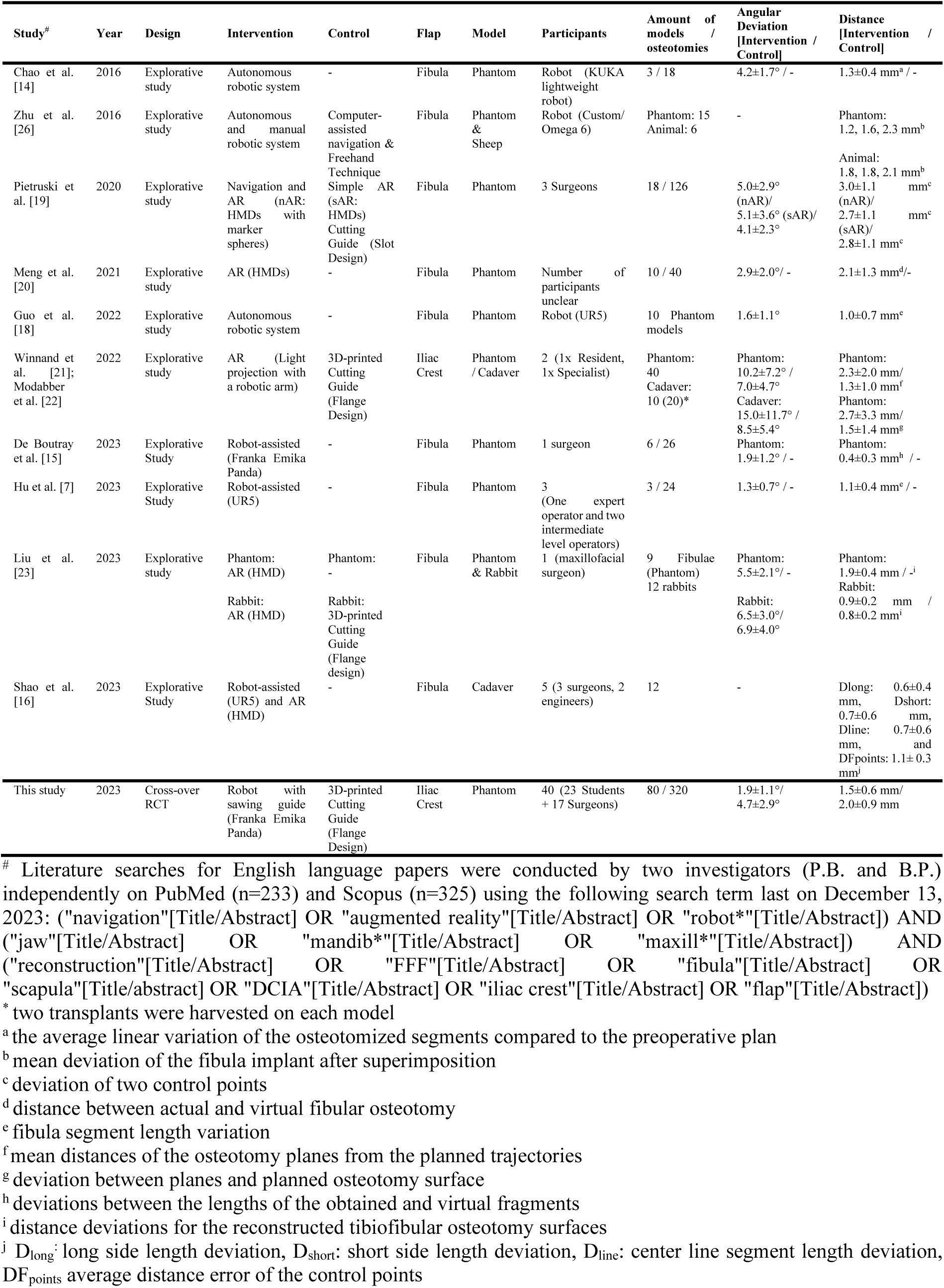

### State of the Art

Hu et al. presented a haptic-guided robotic approach for raising FFFs by sensor-aware hybrid force-motion control [7]. The sensor increased or decreased the motion or stopped the osteotomy when a change in force occurred. Based on the VSP the robotic arm moves to the preplanned trajectory, while the saw can still be controlled by the surgeon. An optical tracker was used to register the position of the fibula and to navigate according to the VSP. In a preclinical study, de Boutray et al. developed a robotic system for FFF raising, where a robotic arm placed a surgical guide with optical tracking markers on the bone which allowed the participants to perform the osteotomies [15]. The collaborative approaches showed angular accuracies of 1.3±0.7° [7] and 1.9±1.2° [15].

In addition to the use of robots with haptic augmentation, systems with visual augmentation have been developed. Pietruski et al. demonstrated an application with AR with head-mounted displays (HMDs) in 2020, comparing a single AR approach to a combined approach with Navigation and AR. 3D-printed cutting guides were used as the control group [19].

In 2021, Meng et al. showed an approach with Mixed Reality using HMDs, guided by voice and gestures [20]. Two other studies used a light projection with a robotic arm of the flap design on the iliac crest instead of a robotic approach in 2022. However, these approaches have shown inaccuracies in visualizing two-dimensional images on a three-dimensional object [21,22].

In 2023, Liu et al. compared AR with HMDs to 3D-printed cutting guides on phantom models of the fibula and on rabbits [23]. Shao et al. investigated a combined approach with AR and robot-assisted navigation [16]. Battaglia et al. presented a workflow for a marker less AR approach with a mobile app that displayed a surgical plan of reconstruction and compared it intraoperatively with the actual anatomy [25].

Besides robot-assisted and AR approaches, Chao et al. investigated the feasibility and accuracy of pre-planned autonomous robotic osteotomies for FFF harvesting. Using VSP, osteotomy planes were generated for three 3D-printed fibula models and programmed into an autonomous robot with a mounted saw [14]. Zhu et al. compared three different methods of FFF harvesting. The first method was an autonomous robotic system with optical tracking, the second was computer-assisted navigation and the third was the freehand technique [26]. Guo et al. conducted a further study about an autonomous robotic system for FFF harvesting, where an algorithm converted the preoperative VSP into motion paths [18]. Accuracies for angular deviations for flap raising with autonomous systems range from 1.6±1.1° to 4.2±1.7°. To the best of our knowledge, autonomous robotic systems have not been tested for DCIA raising [14,26]. Overall, the majority of the studies focused on FFF raising.

Unlike collaborative robots, autonomous systems have higher regulatory requirements of the FDA or MDR [27]. Legal requirements are further increased by the risk of injuring important abdominal structures [28,29] and require patient and surgeon acceptance prior to clinical implementation [30]. While osteotomy angles are not more accurate than those of collaborative approaches, the overall benefit seems to be small [26].

The advantages of collaborative robotic surgery over autonomous systems are consistent with their already established usage in orthopedic surgery [31,32]. Several systems, such as the MAKO or ROSA Knee System, are used for hip and knee replacement, reducing the surgeon’s physical workload while improving the quality, safety and efficiency of osteotomies [33]. While the MAKO system consists of a saw, that is mounted to the robot directly as an end-effector and sets physical limits to protect the cruciate ligaments, the ROSA Knee System places a cutting guide on the surface of the bone, so that the surgeon cuts along the template manually [32,34]. However, flap raising like FFF, DCIA flap or scapula flap are different surgical procedures because not only planes, but a full transplant with soft tissue and most importantly the vascular pedicle is raised. The pedicle is very vulnerable and must be protected during the surgery to prevent flap loss.

Furthermore, the anatomy and especially the vascular supply differs as well. The risk of major bleeding in the knee is significantly lower than the risk of pelvic bleeding from the iliac vessels or abdominal bleeding or infection, which can lead to death [35,36]. As these systems do not have approval for procedures like flap raising or jaw reconstruction, systems like MAKO or ROSA cannot be used one-to-one in OMFS [37].

### Objectives

For these reasons, this study aimed to present a new approach inspired by systems already used in orthopedic surgery [31,32] and preclinical collaborative robot-assisted FFF raising [15]. However, all studies about robot-assisted flap raising in OMFS to date have been exploratory, often lacking gold standard comparisons, while the small number of participants/osteotomies do not adequately account for possible intra- and interrater variability (Table 1).

To address these limitations, we conducted to our knowledge the first prospective, randomized, crossover study to evaluate the feasibility and accuracy of a haptic robot-assisted cutting method compared to conventional 3D-printed cutting guides in DCIA flaps. For this purpose, a static setting with phantom models of the iliac crest was used and DCIA flaps were raised by participants using both methods.

## Methods

### Study design

40 participants with no prior experience in flap raising with 3D-printed cutting guides (medical and dental students, residents or specialists in oral surgery or oral and maxillofacial surgery) were included and performed both methods in a randomized cross-over order (Figure 1). The primary endpoint was the angular deviation of the osteotomy planes between the planned and raised flaps using the robot-assisted method (intervention) and the 3D-printed cutting guide (control). Secondary endpoints were the Hausdorff distance (HD) and average Hausdorff distance (AVD) of the osteotomy planes, the flap raising duration, the perceived workload with NASA-TLX [38] (German version) [39] and the user satisfaction (Figure 2). The carry-over effect as a training effect was considered low since the settings of the two methods were not identical and all participants were novices in iliac crest flap raising.

**Figure 1.**
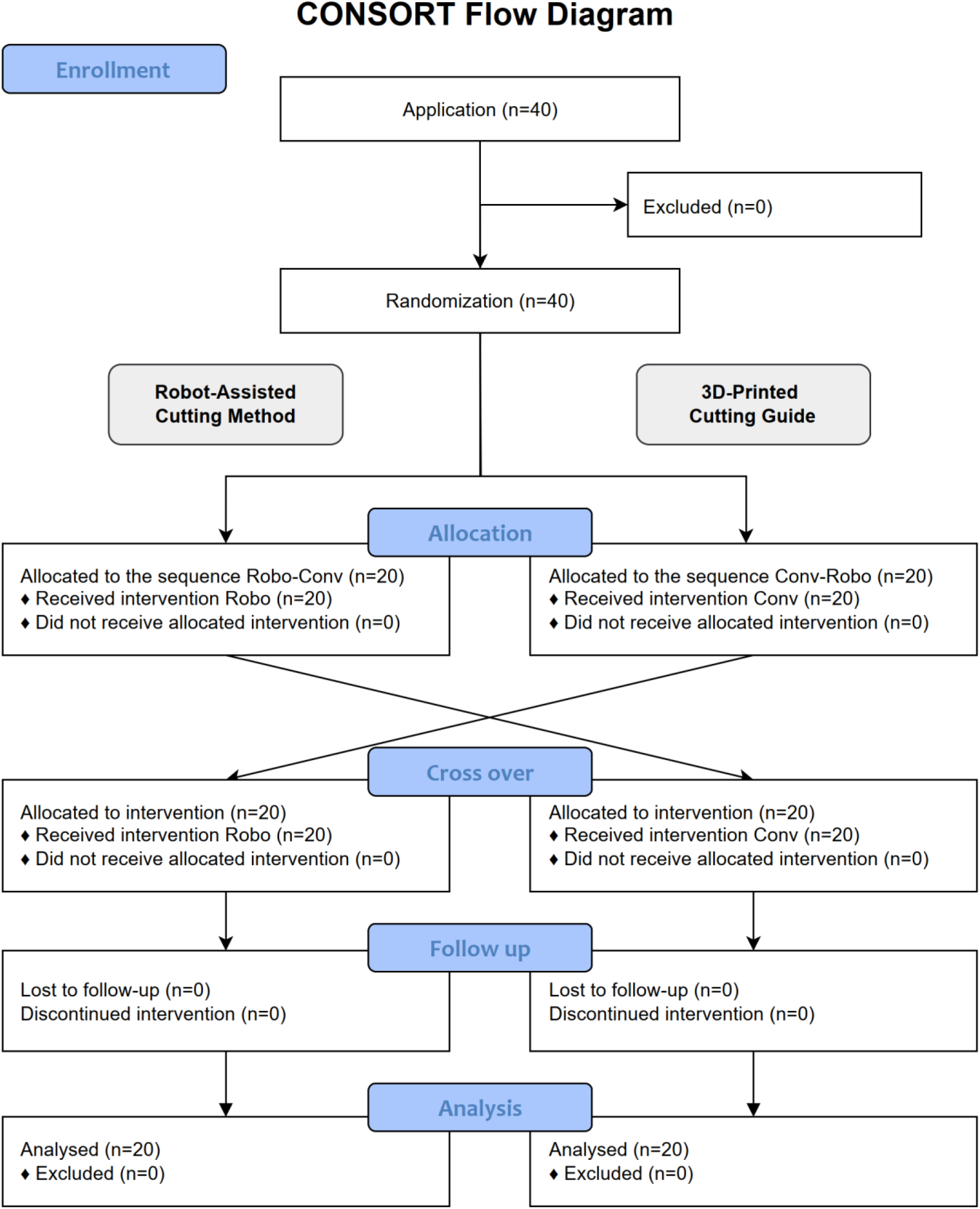
CONSORT flow diagram.

**Figure 2.**
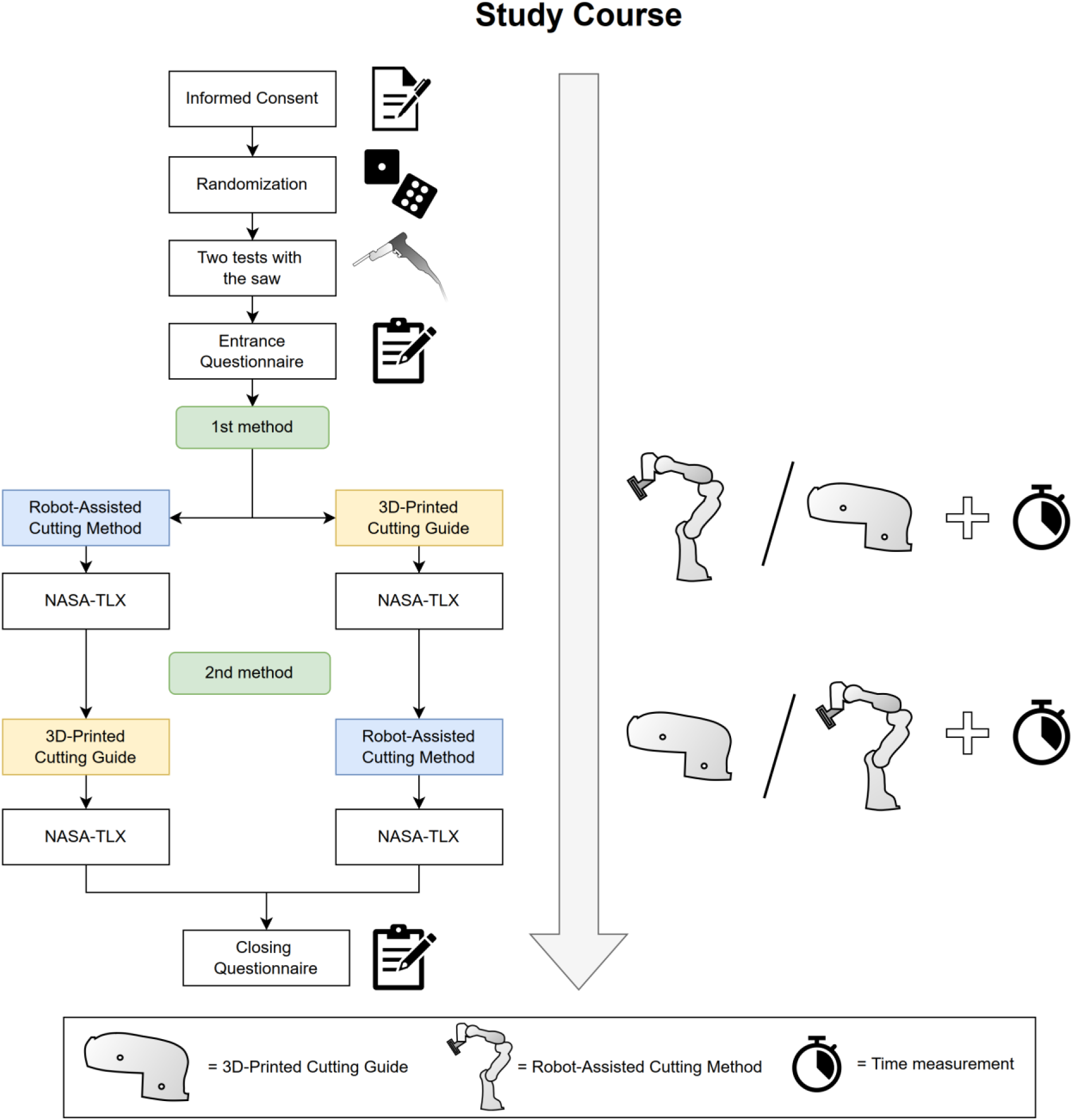
Description of the chronological order of the study.

The study was approved by the Ethics Committee of RWTH Aachen University (approval number EK 23-149, date of approval 20.07.2023) and the study protocol was prospectively registered in the German Clinical Trials Register (DRKS00031358). The study was successfully conducted at the Chair of Medical Engineering of RWTH Aachen University, Germany, from July 31, 2023 to September 21, 2023 and followed the CONSORT 2010 guidelines and its extension for crossover studies [40,41].

### Preparation

For this study, a CT scan was randomly selected from a previous study [42]. After segmentation, the hip model was reduced to the region of the iliac crest and 3D-printed using a Prusa i3 MKS+ (Prusa Research a.s., Prague, Czech Republic) and PLA filament (Beige PLA Filament, made for Prusa, Prusa, Czech Republic) with 0.15 mm layer height and 10% infill.

The software Blender (3.6 LTS, www.blender.org) was used to plan osteotomies by VSP (Figure 3A). Based on the osteotomy planes, the conventional cutting guide for the control group was designed using the displace, solidify and boolean modifiers in Blender. Afterwards, the designed cutting guide was 3D-printed with the same printer model using PETG filament (Prusament, Prusa Research a.s., Prague, Czech Republic) (Figure 4D).

**Figure 3.**
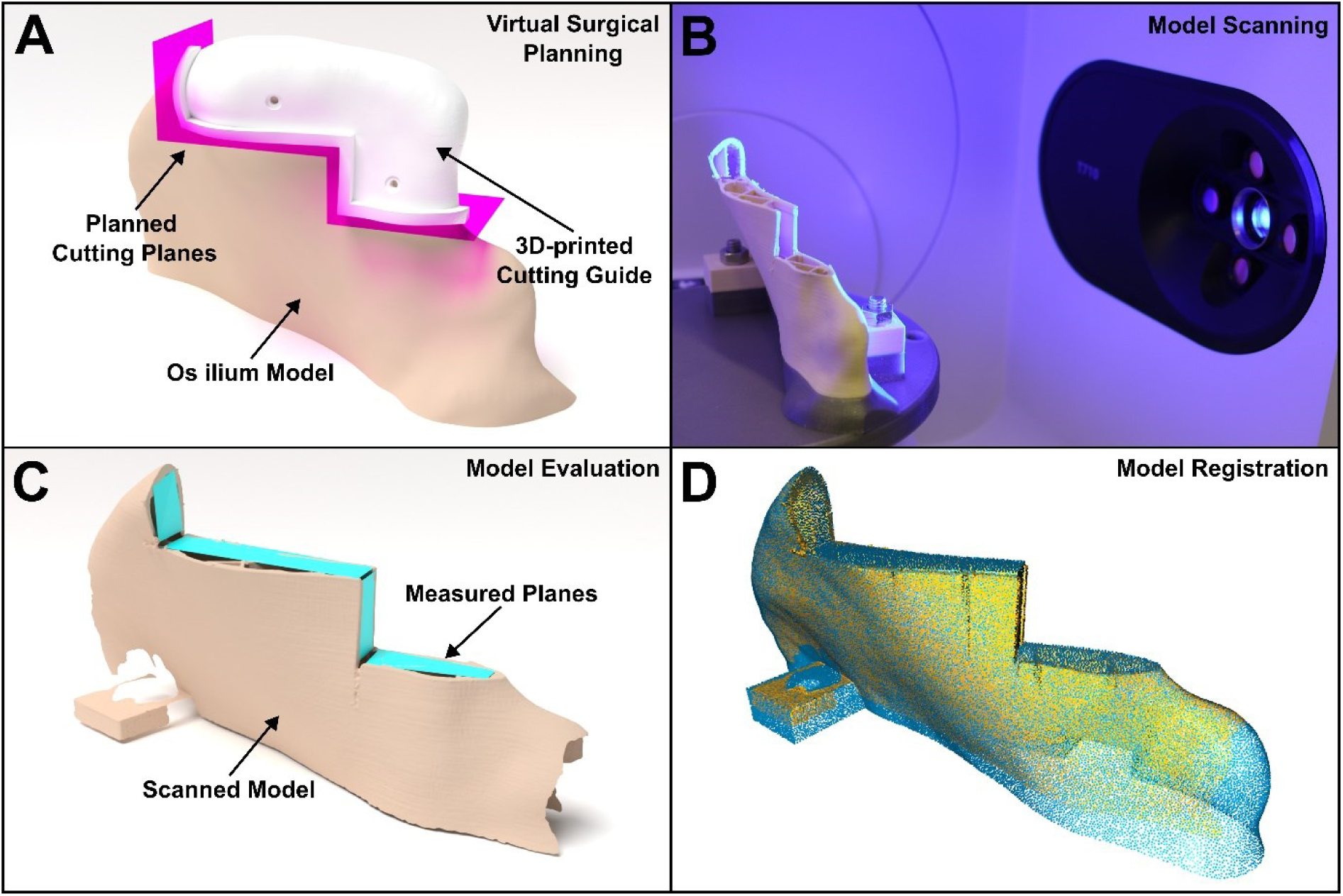
(**A**) Virtual Planning of the transplant and Computer assisted design of the surgical cutting guide in Blender. (**B**) Scanning process of the sawed iliac crest model using a 3D-Scanner (Medit T710, Medit, Seoul, South Korea). (**C**) 3D-Visualization of the scanned model, including planes based on four points, that were used for the evaluation and were created by two independent investigators. (**D**) Iterative closest point (ICP) registration of the scanned model with the planning model.

**Figure 4.**
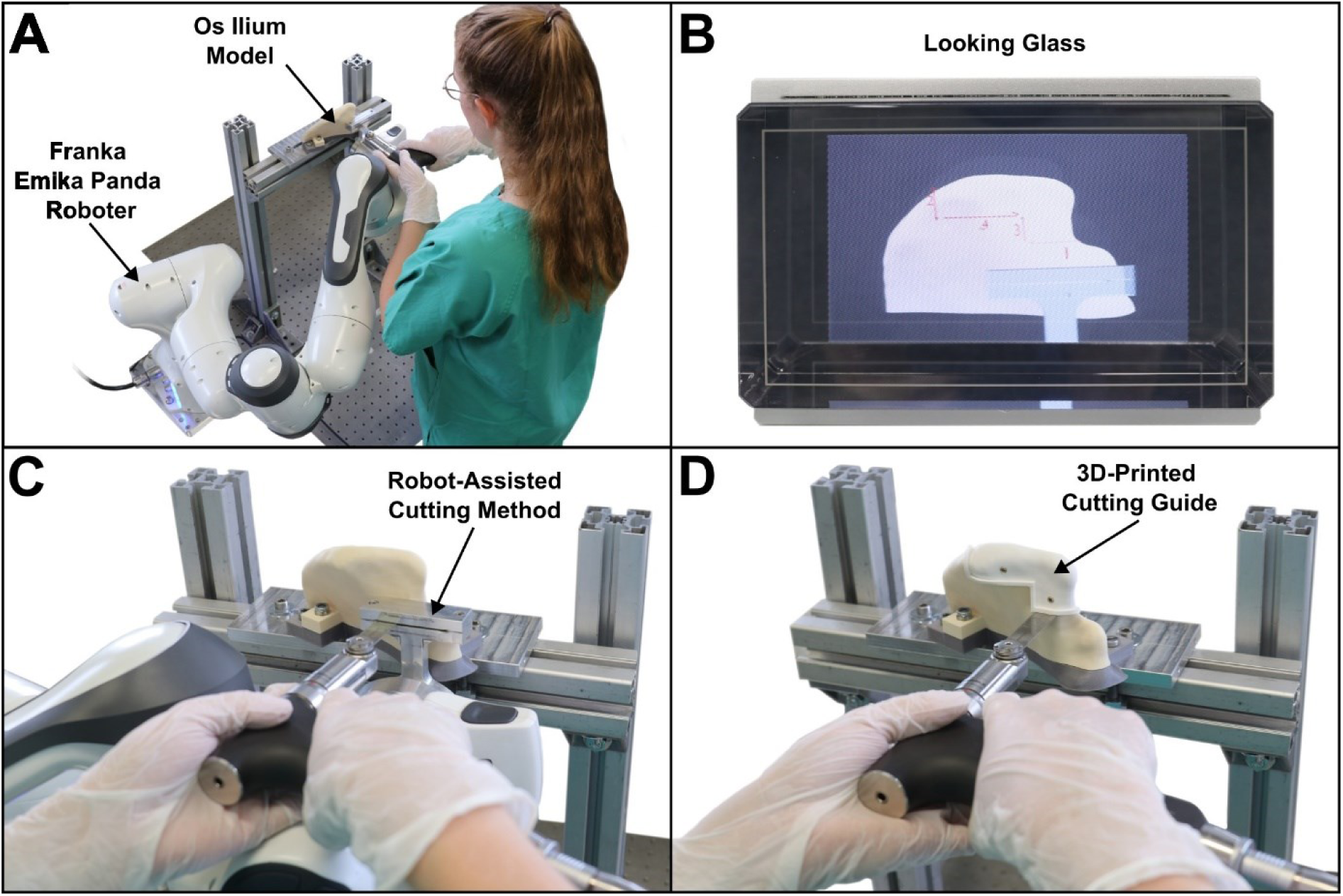
(**A**) A participant performing the osteotomy using the robot-assisted cutting method with the Franka Emika Panda Robot. (**B**) Holographic visualization of the robotic method and of the osteotomy sequence. (**C**) Experimental setup of the robot-assisted cutting method. (**D**) Experimental setup of the conventional 3D-printed cutting guide.

The intervention arm consisted of the robot-assisted cutting method (Figure 4A, C). For this method, a saw guide was first designed in Blender and then fabricated out of Aluminum 7075. The height of the slot was 0.8mm and the depth of the guide was 2cm. The saw guide was mounted on a robotic arm, a Franka Emika Panda (Franka, Munich, Germany).

The Franka Emika Panda was selected due to its seven degrees of freedom, which facilitate a high degree of arm agility. Moreover, the robot exhibits high accuracy, with a position repeatability of ±0.1 mm. Furthermore, the robot is programmable via a multitude of interfaces, including three Franka interfaces, in addition to C++, ROS, ROS2, MATLAB, and Simulink. This makes the robot highly versatile, enabling its application in other specialties. One of the most decisive factors, however, was the robot’s compactness and the internal collision detection mechanism, with a collision detection time of <2ms, which increases patient and user safety [43].

The robot was programmed to place the guide on the surface of the hip model and allowed the participant to perform the osteotomy restricted by the saw guide. The phantom models were mounted on an aluminum frame, which was maintained in a fixed position throughout the duration of the study. To program the right cutting positions, we previously 3D-printed a positioning reference based on the planned CAD/CAM prototype. An additional intermediate position was programmed from which the robot moved to the next osteotomy position. Thereby, the participants still perform the osteotomy themselves and maintain control of the procedure, while the robot positions a saw guide and provides haptic assistance of the osteotomies according to the preprogrammed plan. The specific osteotomy positions for the robotic arm were programmed with ROS (ROS Noetic, Open Source Robotics Foundation) and C++.

Unlike the 3D-printed cutting guide, the shape of the saw guide placed by the robot did not visualize the shape of the flap. Therefore, we implemented a static holographic visualization of the robot-assisted cutting method, to illustrate the dimensions of the flap and the sequence of osteotomies. The iliac crest, osteotomy sequence and saw guide were displayed in the Looking Glass 7.9” (Looking Glass Factory Inc., New York, USA) using the Blender add-on for Looking Glass (Alice/LG, version 2.2).

### Trial

To avoid bias due to a learning effect from previous osseous flap raising with 3D-printed cutting guides, participants with self-performed raising of bone flaps with 3D-printed cutting guides in the past were excluded. Furthermore, left-handed participants were excluded because an adaptive positioning of the robot on the right side was not possible for this study.

Each participant had to fill out written informed consent and an entrance questionnaire before starting the trial. According to a random allocation rule (planned and performed by B.P.) with a balanced block size of 20 for each method, they started either with the 3D-printed cutting guide or robot-assisted cutting method. Before starting the first method, two test planes were sawed on a test block by each participant to get used to the saw (C2 shaver system, Eberle GmbH, Wurmberg, Germany) and the material of the phantom models. After each method, the participants filled out the NASA-TLX score [39]. The duration was measured from the first osteotomy plane to the completion of the last plane. Finally, a closing questionnaire with Likert and open-ended questions was filled out.

### Evaluation

Due to the variability of the raised flaps, scanning in a reproducible manner would be difficult. Instead, we scanned the os ilium models in a standardized manner on a 3D-printed specific mount using a 3D scanner T710 (Medit, Seoul, South Korea) at a resolution of 4 µm (Figure 3B).

The evaluation was performed in a blinded manner by two independent investigators (P.B. and Y.L.). Blinding was performed by an independent person (B.P.). The file names were given random alphanumeric names and all metadata (date of creation, etc.) of the file was removed, as well as any possible identifying content (e.g., labels on the files). Both investigators used Blender to generate planes based on four points, consisting of two triangles for every osteotomy plane (Figure 3C). Each plane was then exported separately as an STL-file. Outliers (planes with a difference of 0.5° or more between the two investigators) were reviewed (B.P.) and were corrected if there were any obvious irregularities (P.B. or Y.L.).

Despite the scanning mount for reproducible scans, during the evaluation of all scanned 80 models, we noted that not all models were perfectly aligned with each other. Therefore, the scans were additionally registered using iterative closest point (ICP) point-to-plane point clouds with the Open3D Python library. Both the originally planned model and the scanned model were registered based on 50,000 points. The registration was run with the following termination parameters: relative_fitness, 1.0×10^−6^; relative_rmse, 1.0×10^−6^; the maximum number of iterations, 100,000. The corresponding transformation matrices (4×4 matrix) were then used to align the created evaluated cutting planes and scanned models to the planned cutting planes.

To evaluate the angular osteotomy plane deviation, the normal vector of the plane was used. Because four points do not necessarily lead to an even plane, the average normal vector of the two triangles of the plane was calculated. Based on the average normal vector and after applying the registration transformation, the angle difference between planned and executed osteotomy planes was measured in degrees. The preoperatively planned angles were set as a reference to 0°. All planes were then automatically calculated using the Trimesh library in Python.

To extract the raised flap, we used the boolean operator with a Python script in Blender according to the reverse engineering principle. Based on the average normal vector, planes were calculated at the same position as the registered osteotomy planes. The exact flap was generated from these planar planes by applying a boolean operator on the complete os ilium model, considering the cutting width of 0.7mm (according to the manufacturer). The volumes of the flaps were calculated in ml. For HD and AVD, the corresponding osteotomy planes from the generated models were used.

All calculations were made using the with the Open3D and Trimesh library in Python.

### Sample Size Calculation and Statistical Analysis

The sample size calculation and statistical analysis were performed in R (version 4.3.0, www.r-project.org). For this purpose, a pretrial was conducted with two medical students and two surgeons, who were randomly assigned to raise DCIA flaps with both methods. Based on the eight harvested flaps, 24 planes were evaluated and used for a simulation-based power analysis for a linear mixed-effects model (LMM) with lmerTest Package [44]. The conventional method had an angular deviation of 4.1±2.1° (mean±sd) and the robot-assisted method 2.1±0.6°. The significance level was set at α=0.05 and the power at 95% resulting in a sample size of 34 subjects. Four more subjects were added to the study to account for dropouts and non-usable data, giving a total of 38 subjects.

The osteotomy angle deviations, the HD and the AVD were also analyzed by LMMs. The dependent variable was the osteotomy angle, while the method (robot-assisted vs 3D-printed) and the orientation of the osteotomy (horizontal vs vertical) were considered independent variables and fixed effects. Mixed effects were the subjects themselves and the osteotomy plane (1-4). The NASA-TLX score was analyzed using a *t*-test, while the duration was analyzed with a Wilcoxon test due to non-normally distributed values. Normal distribution was previously tested using the Shapiro-Wilk test. A p-value <0.05 was considered statistically significant.

## Results

Overall, 40 participants took part in our study. 16 (40.0%) were female and 24 (60.0%) were male. The mean age was 27.1 years (sd 5.3). 17 (42.5%) surgeons and 23 (57.5%) students were included. 16 (40.0%) of the participants were medical students, 7 (17.5%) dental students, 13 residents (32.5%), and 4 specialists (10.0%). The mean study progress was 4.3±0.8 years, while the average years practiced were 5.8±4.5. 38 participants had no previous experience with 3D-printed cutting guides. Two participants were recognized after participation as having some experience with 3D-printed cutting guides. To rule out any possible influence, the analyses were also carried out without them and did not lead to any change in the results of the p-values (Table 2).

**Table 2.** Characteristics of the cohort.

| Parameter |  | Started with Cutting Guide (n=20) | Started with robot-assisted method (n=20) | Total (n=40) |
| --- | --- | --- | --- | --- |
| Sex | Female | 7 (35.0%) | 9 (45.0%) | 16 (40.0%) |
|  | Male | 13 (65.0%) | 11 (55.0%) | 24 (60.0%) |
| Age | Mean (SD) | 25.9 (4.3) | 28.2 (6.1) | 27.1 (5.3) |
| Profession | Student | 13 (65.0%) | 10 (50.0%) | 23 (57.5%) |
|  | Doctor | 7 (35.0%) | 10 (50.0%) | 17 (42.5%) |
| Group | Dental Student | 6 (30.0%) | 1 (5.0%) | 7 (17.5%) |
|  | Medical Student | 7 (35.0%) | 9 (45.0%) | 16 (40.0%) |
|  | Resident | 6 (30.0%) | 7 (35.0%) | 13 (32.5%) |
|  | Specialist | 1 (5.0%) | 3 (15.0%) | 4 (10.0%) |
| Stud Progress (Years) | Mean (SD) | 4.3 (0.9) | 4.4 (0.8) | 4.3 (0.8) |
| Years Practiced | Mean (SD) | 4.9 (2.7) | 6.4 (5.4) | 5.8 (4.5) |
| Previous experience with 3D-printed cutting guides |  | 2 | 0 | 2 (5.0%) |

All in all, 80 models with four osteotomy planes each were evaluated, giving a total of 320 planes. The resulting root mean square error (RMSE) for ICP registration was 0.28±0.05 mm. For the primary endpoint, the robotic-assisted method was with an angular deviation of 1.9±1.1° significantly more accurate, than the 3D-printed cutting guide with an angular deviation of 4.7±2.9° (LMM, p<0.001). Overall, the angular deviation for the robot-assisted cutting method was 2.8° more accurate (Figure 5C). Regardless of the method, the vertical osteotomies showed a lower accuracy of 0.6° (LMM, p=0.008). A visualization of the osteotomy planes for both methods is shown in Figure 5A, B.

**Figure 5.**
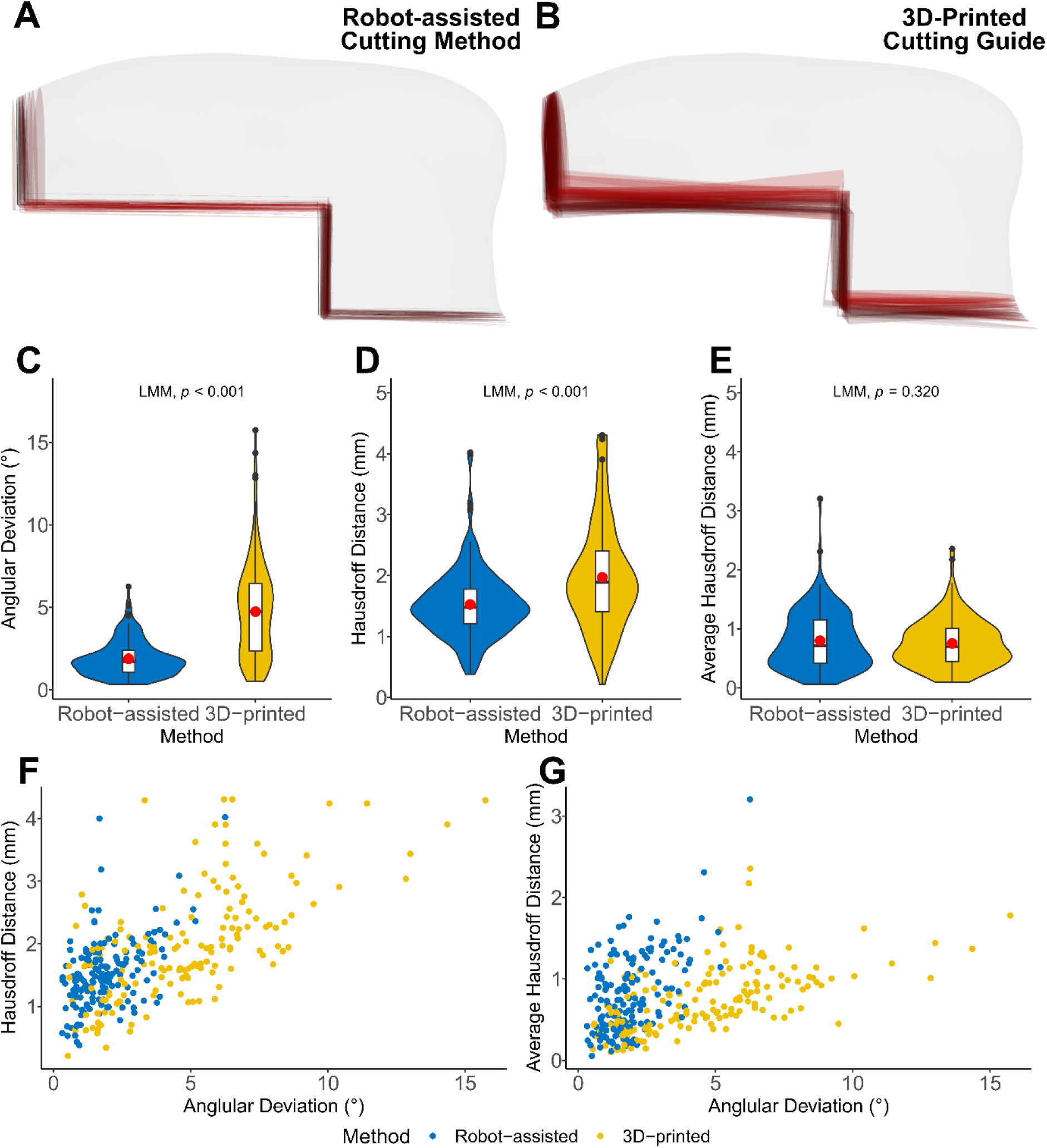
(**A**) Comparison of all 40 osteotomies performed with the robot-assisted cutting method (red) with the planned transplant (white). (**B**) Comparison of all 40 osteotomies performed with the 3D-printed cutting guides (red) with the planned transplant (white). (**C**) Difference of the osteotomy angles between the planned and harvested osteotomy planes for the 3D-printed cutting guide and the robot-assisted cutting method in degrees. (**D**) Hausdorff Distance between the planned and harvested osteotomy planes for the 3D-printed cutting guide and the robot-assisted cutting method in mm. (**E**) Average Hausdorff Distance between the planned and harvested osteotomy planes for the 3D-printed cutting guide and the robot-assisted cutting method in mm. (**F**) Visualization of angular deviation (x-axis) versus HD (y-axis) for both methods. (**G**) Visualization of angular deviation (x-axis) versus AVD (y-axis) for both methods. (**F, G**) Blue points are from the robot-assisted method and yellow points are from the 3D-printed cutting guide.

The HD was 1.5±0.6 mm for the robot-assisted cutting method and 2.0±0.9 mm for the 3D-printed cutting guide (LMM, p<0.001) (Figure 5D). The AVD was 0.8±0.5 mm for the robot-assisted cutting method and 0.8±0.4 mm for the 3D-printed cutting guide (LMM, p=0.320) (Figure 5E). The average volume was 17.32 ml for all raised DCIA flaps and 17.41 ml for the planned DCIA flap.

Subjective workloads were rated significantly lower for the robot-assisted cutting method with an overall score of 38.3±16.5 compared to the conventional method with a total result of 47.7±17.5 (*t*-test, p=0.015) (Figure 6A). The duration was shorter with the 3D-printed cutting guide with 02:07±00:49 min:s compared to 03:14±00:04 min:s for the robot-assisted cutting method (Wilcoxon signed-rank test, p<0.001) (Figure 6B).

**Figure 6.**
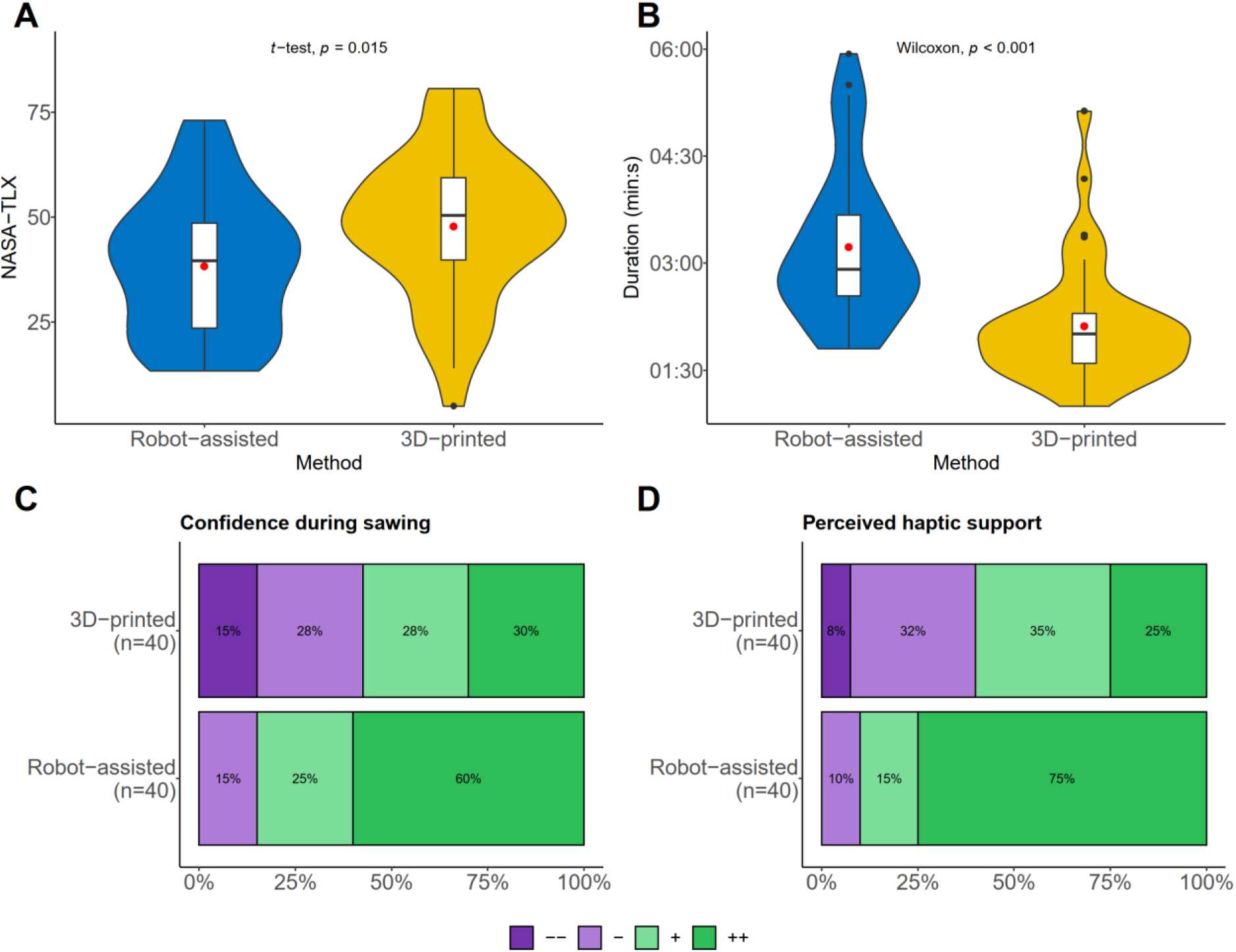
(**A**) Results of the subjectively experienced workload measured with NASA-TLX score using the mean and standard deviation. (**B**) Results of the average duration and standard deviation of the 3D-printed cutting guide and robot-assisted cutting method. (**C**) Results of the second Likert Question about how safe the participant felt during the sawing process, dark purple meaning very poor (1) and dark green meaning very good (4). (**D**) Results for the third Likert Question “The method provided good haptic support during the sawing process”, dark purple meaning very poor (1) and dark green meaning very good (4).

According to the Likert questions (Table 3), the 3D-printed cutting guide was rated to be more intuitive (conventional: 3.4, robot-assisted 3.3, p=0.687) and practical (conventional: 3.4, robot-assisted: 2.8, p=0.008) compared to the robot-assisted method. Both methods were rated equally to be recommended (3.2 for both methods, p=0.975). For the remaining Likert questions, the robot-assisted cutting method was rated superior, especially ratings regarding the accuracy (conventional = 3.1, robot-assisted = 3.5, p=0.033), safety (conventional = 2.7, robot-assisted = 3.4, p=0.001) and haptic support (conventional = 2.8, robot-assisted = 3.6, p<0.001) were significantly better (Figure 6C, D).

**Table 3.** Likert questionnaires.

| Questions | 3D-printed (n=40) | Robot-assisted (n=40) | Total (n=80) | p value |
| --- | --- | --- | --- | --- |
| The method helps to precisely implement the planned osteotomy. | 3.1 (0.8) | 3.5 (0.6) | 3.3 (0.7) | 0.033 |
| I felt safer when sawing with the method. | 2.7 (1.1) | 3.4 (0.7) | 3.1 (1.0) | 0.001 |
| The method provided good haptic support. | 2.8 (0.9) | 3.6 (0.7) | 3.2 (0.9) | < 0.001 |
| The method is intuitive. | 3.4 (0.7) | 3.3 (0.7) | 3.4 (0.7) | 0.687 |
| The method is easy to use. | 3.3 (0.9) | 3.5 (0.6) | 3.4 (0.8) | 0.617 |
| The method helps to saw effectively. | 3.2 (0.9) | 3.4 (0.7) | 3.3 (0.8) | 0.551 |
| The use of the method increases patient safety. | 3.2 (0.9) | 3.4 (0.7) | 3.3 (0.8) | 0.511 |
| The method improves the outcome of flap harvesting. | 3.2 (0.9) | 3.4 (0.8) | 3.3 (0.8) | 0.551 |
| I would recommend the use of the method. | 3.2 (0.8) | 3.2 (0.8) | 3.2 (0.8) | 0.975 |
| In my opinion, the method is practical. | 3.4 (0.7) | 2.8 (0.9) | 3.1 (0.8) | 0.008 |

The open questions revealed the following: Many participants mentioned the good haptic guidance and accuracy of the robot-assisted cutting method, especially for beginners. However, some participants also mentioned a limited view of the bone due to the mounted saw guide as a negative aspect. Regarding the 3D-printed cutting guide, lower haptic guidance and the time-consuming fixation with screws were criticized. Positive aspects were easy handling and the visualization of the transplant by the shape of the cutting guide (Table 4).

**Table 4.**
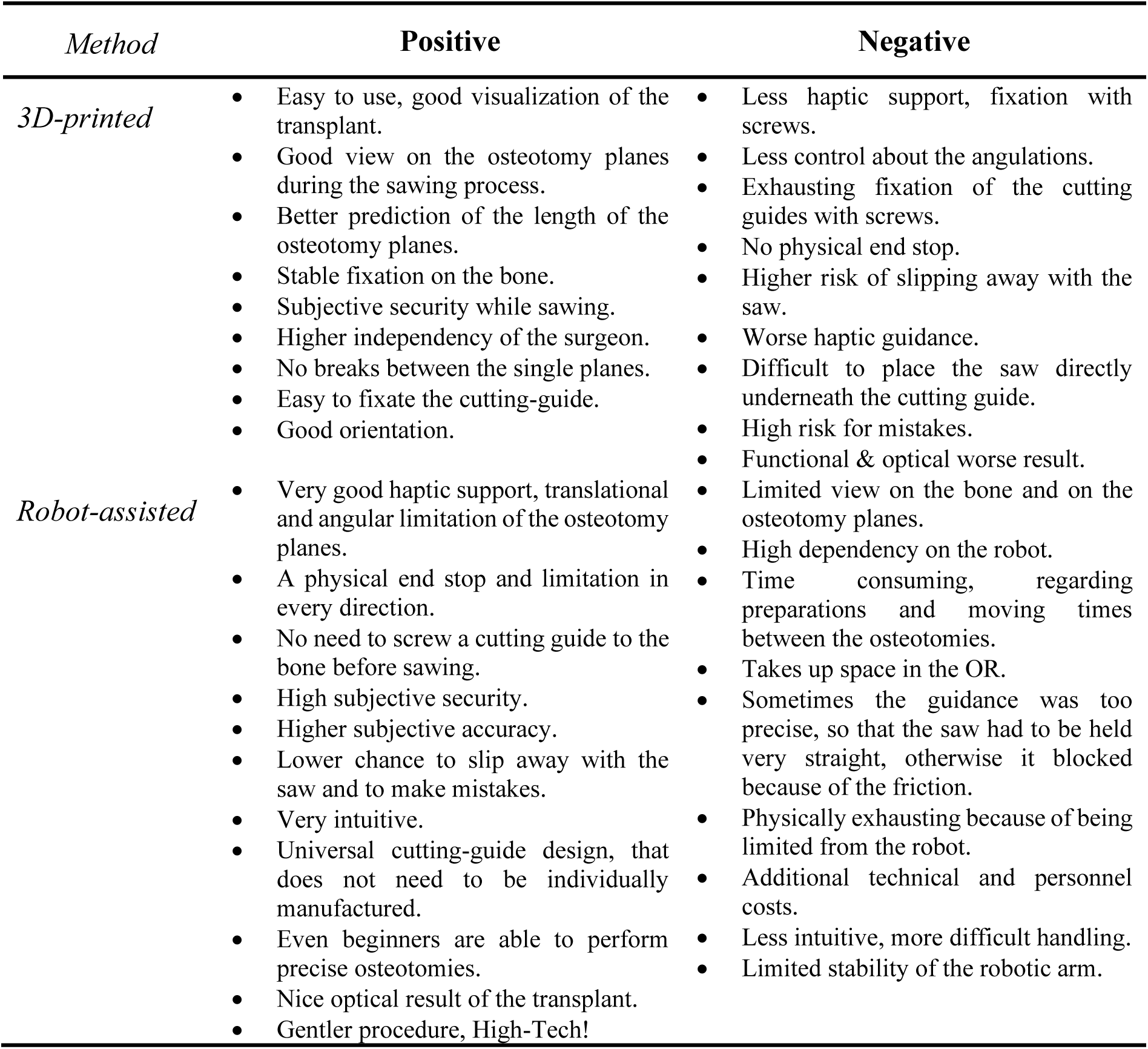

| *Method* | **Positive** | **Negative** |
| --- | --- | --- |
| *3D-printed* | • Easy to use, good visualization of the transplant.<br>• Good view on the osteotomy planes during the sawing process.<br>• Better prediction of the length of the osteotomy planes.<br>• Stable fixation on the bone.<br>• Subjective security while sawing.<br>• Higher independency of the surgeon.<br>• No breaks between the single planes.<br>• Easy to fixate the cutting-guide.<br>• Good orientation. | • Less haptic support, fixation with screws.<br>• Less control about the angulations.<br>• Exhausting fixation of the cutting guides with screws.<br>• No physical end stop.<br>• Higher risk of slipping away with the saw.<br>• Worse haptic guidance.<br>• Difficult to place the saw directly underneath the cutting guide.<br>• High risk for mistakes.<br>• Functional & optical worse result. |
| *Robot-assisted* | • Very good haptic support, translational and angular limitation of the osteotomy planes.<br>• A physical end stop and limitation in every direction.<br>• No need to screw a cutting guide to the bone before sawing.<br>• High subjective security.<br>• Higher subjective accuracy.<br>• Lower chance to slip away with the saw and to make mistakes.<br>• Very intuitive.<br>• Universal cutting-guide design, that does not need to be individually manufactured.<br>• Even beginners are able to perform precise osteotomies.<br>• Nice optical result of the transplant.<br>• Gentler procedure, High-Tech! | • Limited view on the bone and on the osteotomy planes.<br>• High dependency on the robot.<br>• Time consuming, regarding preparations and moving times between the osteotomies.<br>• Takes up space in the OR.<br>• Sometimes the guidance was too precise, so that the saw had to be held very straight, otherwise it blocked because of the friction.<br>• Physically exhausting because of being limited from the robot.<br>• Additional technical and personnel costs.<br>• Less intuitive, more difficult handling.<br>• Limited stability of the robotic arm. |

In total, 21 participants preferred the robot-assisted cutting method, and 19 participants preferred the 3D-printed cutting guides.

## Discussion

To our knowledge, this is the first RCT to compare a collaborative robot-assisted cutting method with 3D-printed cutting guides and to demonstrate its feasibility for raising DCIA flaps. The main findings were a higher angular accuracy and a reduction of the subjective workload of the robot-assisted cutting method compared to 3D-printed cutting guides. With less than four minutes, both methods were sufficiently fast. The HD was also lower for the robot-assisted method, while the AVD showed no significant difference.

On average, the robot-assisted cutting method (1.9±1.1°) was 2.8° more accurate than the conventional 3D-printed cutting guides (4.7±2.9°). These findings are comparable to the results from the studies conducted by Hu et al. [7] and de Boutray et al. [15] with angular deviations of 1.3±0.7° respectively 1.9±1.2°. Common numbers for angular deviations for 3D-printed cutting guides are 4.1±2.3°, 7.0±4.7°, 8.5±5.4° and 6.9±4.0° (Table 1). The results show that the preoperative plan (CAD/CAM) is accurately transferred to the surgical site by the robot-assisted cutting method.

Nevertheless, the previously described robot-assisted methods for FFF raising had a purely exploratory design and only a few participants were included [7,15]. Consequently, the inter-rater variance was not considered and not all, but many other studies had no control group [7,14–16,20]. In contrast [18], our study is a confirmatory study, including study registration with sample size calculation including a large number of participants and comparison to the gold standard (3D-printed cutting guides) as a control. This is however necessary to evaluate the effectiveness of the method and to attribute causality [45].

Interestingly, there was no difference in translational error (for AVD) between the two methods. This suggests that there was mainly a rotational error of the osteotomy depending on the method (Figure 5F). The AVD results in many very low values (Figure 5G) due to the crossing of the performed osteotomy with the planned osteotomy. A parallel translation would have resulted in significantly higher AVD values. The AVD values of this study are comparable to the translational error of 1.2 mm found in the study conducted by Zhu et al [26].

The angular deviation should be considered in the context of the lack of a standardized design for 3D-printed cutting guides. In this regard, slot and flange designs are the common ways to guide the surgeon during osteotomy [46,47]. Slotted guides have a smaller range of motion (depending on their design) because they constrain the saw to more dimensions, which may explain the observed differences between the robot-assisted and 3D-printed guide methods. However, in the study by Pietruski et al, the 3D-printed cutting guide with a slot design for FFF raising showed an angular deviation of 4.1±2.3° [19], which is comparable to our flange-designed 3D-printed cutting guides. In addition to the choice of slots or flanges, the length of blade guidance is also critical. Usually, an increased depth of the guide leads to higher guidance. While the guide for the robot-assisted cutting method had a depth of 2 cm, the depth of the flange of the 3D-printed cutting guide was only 5mm.

It is important to note that as DCIA flaps require connected osteotomies to raise the flap, a total slot design for all osteotomies is not an option, while the slots themselves already lead to a larger, more invasive cutting guide. Therefore, many studies used a flange design for 3D-printed cutting guides to raise DCIA flaps in real clinical cases [48–51]. Some studies partially designed a guide with slots, however only the vertical osteotomies were performed through the slots [47,52] (Table 1). The accuracy of cutting guides is further affected by the position of the guide on the bone and by the fixation with screws which could explain the lower accuracy of cutting guides compared to the robot-assisted method.

When evaluating the clinical relevance of the accuracies of osteotomies and surgical cutting guides in our study and the literature, the reproducibility and comparability of those are limited. Besides different cutting guide designs, there are also different methods to evaluate the accuracy of CAS. Landmarks, superimposition and resection planes are possible ways for evaluation [53], while the image quality and the segmentation itself also influence the subsequent steps of the CAS [54].

Besides these technical considerations, the primary goal of maxillofacial reconstruction is aesthetic and functional restoration of the jaw. One of the most important functional outcomes is the dental occlusion, as patients are sensitive to even the smallest changes [55]. In addition to changes in dental occlusion, mandibular deviation can be caused by changes in the position of the condyles [56]. Unlike the upper jaw, the mandible has many muscles attached to it, which further affect the position of the jaw through muscle tone [57]. Furthermore, dental rehabilitation is highly dependent on an accurate reconstruction of the jaw [58]. However, no numbers are available to quantify a desired outcome in terms of angular or linear deviation of the reconstructed jaw [59].

The outcome of the reconstruction depends on several factors, including the translational and rotational errors of the osteotomies of both the jaw resection, and the raised bone flap. Both angles accumulate to the overall margin of error, not only in translation but also in rotation of the mandible [60], which will cause corresponding inaccuracies in the position of the condyles in the temporomandibular joint, the contour of the jaw, or the dental occlusions (Figure 7 A, B). Figures 7C and D illustrate the potential error caused by osteotomy angle deviations of 2° and 5° of a raised flap, showing an increased distance between the two condyles and therefore a change in condyle position in the articular fossa. This suggests that a 2.8° angular deviation of the osteotomy (planned vs. performed) may have a major impact, with accurate reconstruction being critical to multiple rehabilitation factors of the patient, whereas translation errors of approximately 1 mm reported by others and by us seem to contribute less. Current angular accuracies of mandibular reconstruction with classical CAS range from 0.9°-17.5° and linear deviations range from 0-12.5mm using condylar measurements, indicating that there is still an issue here [53].

**Figure 7.**
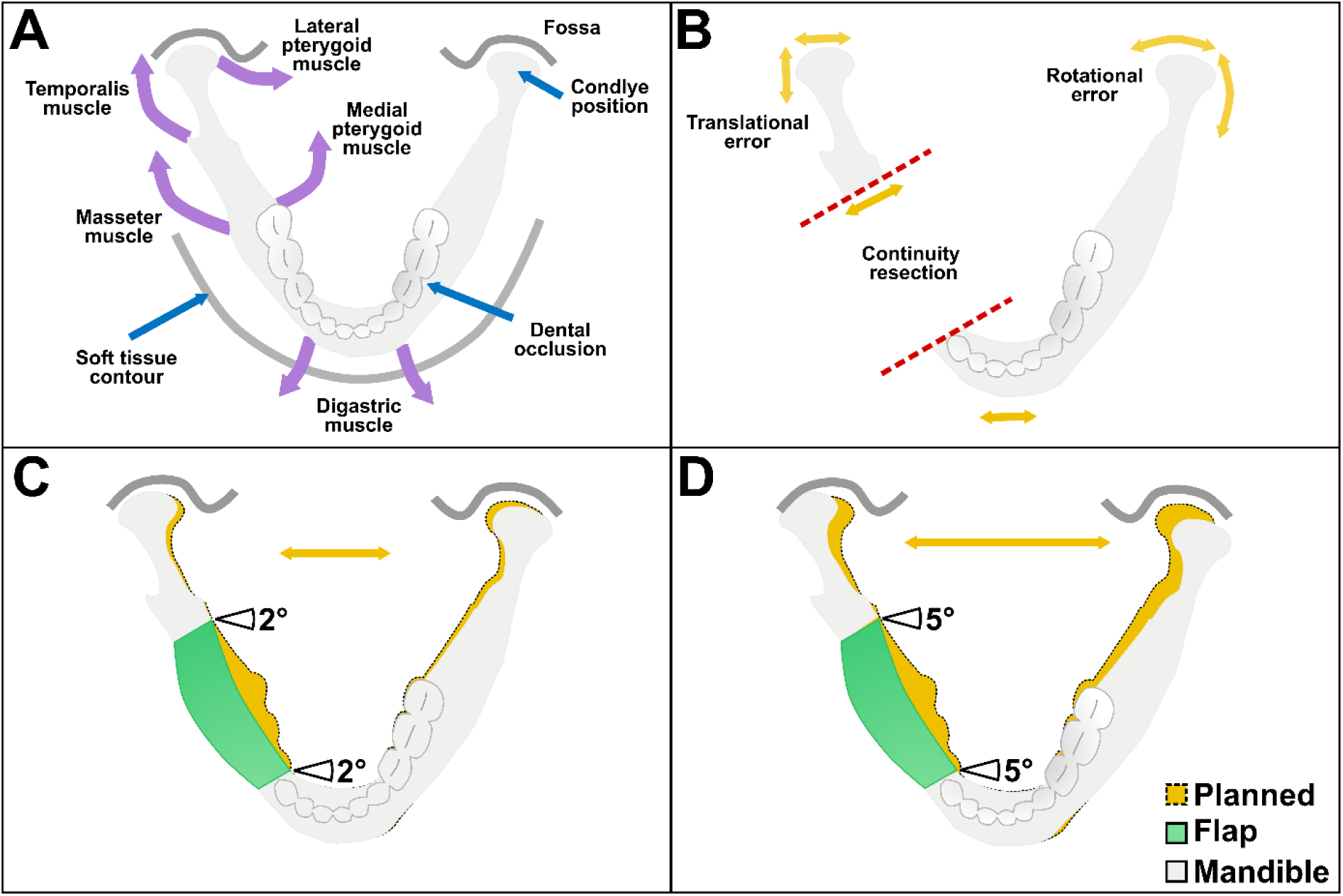
(**A**) Schematic visualization of muscles (purple arrows: digastric, masseter, temporalis, lateral and medial pterygoid muscle) affecting functional outcomes (blue arrows: condyle position in the articular fossa, dental occlusion and soft tissue contour) of the mandible. (**B**) Possible sources of error (yellow arrows: translation and rotation) in accuracy and functional results during mandibular reconstruction. Red dashed line the conducted discontinuity resection. (**C**) Simulated translational error of the condyle position with an angular deviation of the osteotomy angles of 2°. (**D**) Simulated translational error of the condyle position with an angular deviation of the osteotomy angles of 5°.

Nevertheless, participants reported that the 3D-printed cutting guide was more practical and intuitive. 3D-printed cutting guides have been used for decades now and were tested in multiple scenarios [61]. They were originally introduced by Radermacher et al. at the Helmholtz Institute for Biomedical Engineering at RWTH Aachen University in the early 1990s [62,63]. Their design is easy to understand, as it shows the shape of the flap to be harvested.

The limited view of the osteotomy caused by the guide mounted onto the robotic arm could easily be improved by increasing the distance between the bone and the cutting guide. Attaching the saw directly to the robotic arm like the MAKO robot [32] might further improve depth control and protect the abdomen. With trials in a more clinical environment, the usability of the robot-assisted cutting guides could be improved further regarding the handling and placement in the operating room.

However, surgeons are already relieved from physically exhausting tasks and can focus on the precise execution of flap harvesting [64], which is also consistent with the reported subjective workload values in our study. Combined with optical tracking, the robot-assisted cutting method would not require patient specific manufacturing and could be adapted during surgery. This would increase the intraoperative flexibility and overcome the disadvantages of conventional cutting guides, such as high production costs and longer preoperative lead times [15]. Furthermore, invasive fixation of the guide with screws would no longer be necessary.

Collaborative robotic systems either place a physical guide and/or use VSP to transfer the osteotomy planes to the robot. Thereby the osteotomy angle is pre-set by the robotic arm [15]. More inexperienced surgeons could profit from the limited degree of freedom for the sawing blade provided by the saw guide, as there is a lower risk of accidentally slipping away with the saw.

In addition to that, the robot-assisted method also provides a physical end stop, that can protect the pedicle and other abdominal soft tissue behind the iliac crest from being harmed. This is particularly important for raising DCIA flaps, as injuries of the abdominal cavity can lead to potentially lethal consequences [35].

Compared to an autonomous system performing the osteotomy, a collaborative approach has the advantage that the surgeon is always in control, which could lead to better acceptance by both patients and surgeons and facilitate translation from a regulatory perspective. DCIA flap raising is much more complex than FFF raising because it requires the osteotomy of a combination of linked planes. This implies that the individual osteotomies need to be coordinated not only in terms of angular deviation but also regarding length and distance.

Robot-assisted flap raising in OMFS still requires further investigation and interdisciplinary research including surgeons, technicians, and industry, to improve the application of robot-assisted cutting methods during a realistic procedure. Cadaver studies, haptic guidance and real-time navigation could create new findings for reconstructive surgeries. In complex situations, osteotomies may be performed using robot-assisted Er:YAG laser [65]. In combination with planning algorithms or Artificial Intelligence, the transplant could be planned and programmed into the robot [18].

Nevertheless, there are some limitations of our study. First, we only assessed both methods in a static setting. During a real surgical procedure, optical tracking combined would be necessary [15]. Dynamic motion control could increase the stiffness of the robotic arm and thereby reduce errors caused by the movement of the robot.

Furthermore, the lack of clinical results was also caused by using phantom models in an artificial setting, without surrounding structures such as soft tissue and without having to consider the limited space in the surgical field. However, the use of the same phantom model, with the same transplant planning in an identical setting allowed a standardized study for reliable results regarding inference statistics.

Since the study aimed to assess the general feasibility of the system for raising DCIA flaps, a next step to advance clinical translation should be to investigate the system in a dynamic setting such as cadavers. As in the operating room, the entire body will be present, and optical tracking with navigation systems will be performed. This would be a more realistic scenario, which first simulates how the robotic arm acts in the surgical situs and second investigates the feasibility of the system regarding limited space in the operating room.

In experimental and static settings, the haptic robot-assisted method is a good alternative to 3D-printed cutting guides for raising DCIA flaps. The increased angular accuracy obtained with the robot-assisted method for DCIA harvesting is comparable to the current outcomes of preclinical studies on robotic methods used for FFF harvesting. Furthermore, robotic approaches can prevent the need for invasive fixation of a 3D-printed cutting guide and allow for intraoperative planning and flexible planning adaptation. The flange design of the 3D-printed gutting guide resulted in a higher rotational error, but only in a small translational error, which was comparable for both methods. To verify these outcomes, the next step will be to test the results in a dynamic environment with a moving phantom or a cadaver.

## Acknowledgments

We would like to thank the Institute of Anatomy of the RWTH Aachen University and Mr. Heiko Löffler from the Eberle company for providing us the oscillating saws and for the technical support during the study. We would like to thank Mr. Adalbert Mazur from the Scientific Workshop of the Medical Faculty of RWTH Aachen University for producing the sawing guide.

## Declaration

### Author Contributions

Conceptualization, B.P., P.B., R.R. and F.H.; methodology, P.B., B.P., S.D., M.F. and J.E.; software, P.B., B.P. and R.R.; validation, P.B., Y.L., B.P., M.F. and K.X.; formal analysis, P.B., B.P., K.R. and A.R.; investigation, P.B. and B.P.; resources, K.R., F.H., R.R. and B.P.; data curation, P.B. and Y.L.; writing— original draft preparation, P.B.; writing—review and editing, B.P., P.B., Y.L., S.D., J.E., K.X., A.R., K.R., R.R., M.F. and F.H.; visualization, P.B. and B.P.; supervision, B.P.; project administration, B.P.; funding acquisition, B.P.; All authors have read and agreed to the published version of the manuscript.

### Data Availability Statement

The data presented in this study are available on request from the corresponding author.

### Funding

Behrus Puladi was funded by the Medical Faculty of RWTH Aachen University as part of the Clinician Scientist Program. We acknowledge FWF enFaced 2.0 [KLI 1044, https://enfaced2.ikim.nrw/] and KITE (Plattform für KI-Translation Essen) from the REACT-EU initiative [https://kite.ikim.nrw/, EFRE-0801977].

### Institutional Review Board Statement

The study has been approved by the Institutional Review Board (or Ethics Committee) of University Hospital RWTH Aachen (protocol code EK 23-149 and date of approval: 20.07.2023).

### Informed Consent Statement

Informed consent was obtained from all subjects involved in the study. Figure 3A shows the first author, who consented to have her photograph included in Figure 3.

### Competing Interests

The authors declare no competing interests.

## Notes

### Competing Interest Statement

The authors have declared no competing interest.

### Clinical Trial

DRKS00031358

### Clinical Protocols

https://drks.de/search/de/trial/DRKS00031358

### Author Declarations

Ethics Committee of RWTH Aachen University (approval number EK 23-149, date of approval 20.07.2023)

